# Human resources for health, service coverage and maternal and perinatal outcomes in Benin, Malawi, Tanzania and Uganda

**DOI:** 10.1101/2024.07.02.24309845

**Authors:** Ann-Beth Moller, Joanne Welsh, Max Petzold, Amani Siyam

## Abstract

A well-performing and competent health workforce (HWF) is at the core of health systems yet many countries are facing a human resources for health (HRH) crisis. A prerequisite for achieving universal health coverage, including fulfilling the Sustainable Development Goals related to women and newborns, is an adequate supply of health workers able to provide quality of care at all levels of the health system. Thus, we evaluated how HRH policies and strategies influenced trends of maternal and newborn workforce densities and assessed the association between HWF densities, service coverage and health outcomes in Benin, Malawi, Tanzania and Uganda. We applied the READ framework (Ready your materials; Extract data; Analyse data and Distil findings) for our HRH policy and strategy document analysis and conducted a comparative analysis including three HWF densities (medical doctors, nursing and midwifery personnel) two health services, and five health outcome variables. Twenty HRH policies and strategies were included in the analysis published from 2010 to 2021. The scope of the HRH policies and strategies were described in four dimensions; availability, accessibility, acceptability and quality. We found that all policies and strategies addressed aspects related to availability and accessibility as well as the need for HRH quality improvements whereas acceptability was poorly represented. The comparative analysis revealed that service coverage and health outcomes appear to be insensitive to the fluctuations in HWF densities and related HRH policies as very little or no reduction was seen in outcomes from 2010 to 2020. There is a need to tackle the availability, accessibility, acceptability and quality of the HWF. Evidence needs to be translated into policy and practice otherwise the HWF in these countries will continue to struggle, affecting progress and realizing womens’ and newborn’s human rights to health.

## Background

Many countries are facing a human resources for health (HRH) crisis, lacking a competent health workforce (HWF) required to deliver universal health coverage (UHC) matching population needs [1], including care for women and newborns as targeted by the Sustainable Development Goals (SDG) and the UHC agenda [2, 3]. The importance of the HWF in achieving SDG goal 3 “ensure healthy lives and promote well-being for all at all ages” is reflected in SDG target 3.c “substantially increase health financing and the recruitment, development, training and retention of the HWF in developing countries, especially in least developed countries and small island developing States” and indicator 3.c.1 “health workforce density and distribution”. [2] Sub-Saharan Africa (SSA) is confronted with the lowest HWF densities with 2.3 medical doctors, and 12.6 nursing and midwifery personnel per 10 000 population, significantly below the global level of 16.3 medical doctors, and 39.4 nursing and midwifery personnel per 10 000 population. [4] Projections suggest that by 2030 the global HWF shortage will approximate to ten million. SSA will be challenged with the highest shortage of 5.3 million which equates to 52% of the global projected shortage by 2030. [1] The shortage is often linked to lack of investment in education, an absence of HRH policies, international migration of health workers and a weak information system monitoring the stock of the labour market dynamics to inform policies and programmes. [5] The COVID-19 pandemic exacerbated health care worker shortages and highlighted the fragility of the health care system. Those working during the pandemic were faced with risk of infection, exhaustion, stress, and anxiety which combined, led to compromised physical and mental health for numerous health workers, with some leaving their profession. [6–9]

In addition to HRH shortages, evidence suggests that prior to entering the HWF, pre-service training does not always meet international standards. [10] The knock-on effect is reduced competence levels [10, 11] which, in turn, affects quality of care delivered. Furthermore, inadequate remuneration packages and lack of an enabling working environment can leave health care workers demotivated and unprepared to deliver care required. [12–14] Collectively, these factors influence health outcomes for the population. Maternal and newborn health (MNH) is one example. Under SDG 3 and the Global Strategy for Women’s, Children’s and Adolescents’ Health (2016–2030) there are specific targets related to MNH. [2, 15] These include ensuring women have access to family planning, antenatal care, and increasing the number of births attended by a skilled health personnel, also known as skilled birth attendant (SBA). [16, 17] As a result, essential services such as antenatal, intrapartum and postnatal care have been more and more accessible with evidence suggesting increased coverage for all these services in SSA. [18] Access to skilled health personnel during childbirth, a prominent indicator in the SDG framework (SDG indicator 3.1.2), increased globally from 81 to 86 % between 2015 and 2022 with SSA experiencing the lowest coverage (70%) in 2022. [4] Despite this, high levels of maternal and newborn morbidity and mortality still persist. [4]

In 2021 an estimated 1.9 million newborns were stillborn at 28 weeks or more of gestation with SSA accounting for almost half (45%) of these stillbirths. [19] Latest preterm birth estimates suggest stagnation in SSA as 10.1% of all livebirths (95% credible interval (CrI): 8.5-12.6%)) in 2020 were born preterm which shows no change from the 10.1% (95% CrI: 8.6-12.7%) in 2010. [20] Lack of progress is also seen for low birthweight (LBW) in SSA, which has the second highest LBW prevalence after southern Asia in 2020 (24.4% (95% CrI: 20.8-29.3)), with an estimated prevalence of 13.9% (95% CrI: 12.4-15.7%) in 2020 compared to 15.7% (95% CrI: 14.-17.7%) in 2000. [21]

Preterm birth and LBW are not only associated with greater risk of mortality but also greater risk of short- and long-term morbidities throughout the life course, with preterm birth being the main risk factor for death in children below five years, accounting for approximately 18% of such deaths. [22] Furthermore, preterm and LBW births hugely affect pregnant people, families, society and economies. [23–26]

The World Health Organization (WHO) prominently positioned a well-performing and competent HWF as one of the six building blocks for health system strengthening. [27] To ensure the well-being of women and newborns, as well as positive health outcomes for both groups, women and newborns need access to a health system which provides timely quality care, delivered by educated and competent providers. However, many countries including many in SSA struggle to ensure the four essential dimensions for healthcare services defined as: Availability, Accessibility, Acceptability and Quality also referred to as the AAAQ Framework. [28] Box 1. provides the operational definition of the AAAQ framework dimensions.

### Box 1.

**Availability, Accessibility, Acceptability and Quality (AAAQ) framework**

**Box 1. Availability, Accessibility, Acceptability and Quality (AAAQ) framework**
| Framework dimensions | Operational definition |
| --- | --- |
| Availability | The sufficient supply, appropriate stock of health workers, with the relevant competencies and skill mix that corresponds to the health needs of the population. |
| Accessibility | The equitable distribution of health workers in terms of travel time and transport (spatial), opening hours and corresponding workforce attendance (temporal), the infrastructure's attributes (physical—such as disabled-friendly buildings), referral mechanisms (organizational) and the direct and indirect cost of services, both formal and informal (financial). |
| Acceptability | The characteristics and ability of the workforce to treat all patients with dignity, create trust and enable or promote demand for services; this may take different |
|  | forms such as a same-sex provider or a provider who understands and speaks one's language and whose behaviour is respectful according to age, religion, social, cultural values, etc. |
| Quality | The competencies, skills, knowledge and behaviour of the health worker as assessed according to professional norms (or other guiding standards) and as perceived by users. |

Source: Campbell J, Dussault G, Buchan J, Pozo-Martin F, Guerra Arias M, Leone C, Siyam A, Cometto G. A Universal Truth: No Health Without a Workforce: Third Global Forum on Human Resources for Health Report. Geneva : Global Health Workforce Alliance and World Health Organization; 2013.

Gaps in these dimensions have led to a stagnation in outcomes related to MNH [19, 20, 29–31], and jeopardize global efforts to implement and meet targets included in various strategies such as Ending Preventable Maternal Mortality [16], Every Newborn Action Plan [17] , Global Strategy for Women’s, Children’s and Adolescents Health 2016-2030 [15] and targets included in the SDG agenda [2]. To ensure improvement in MNH outcomes it is important to understand how national HRH, and other national policies and strategies have influenced MNH outcomes and related health service to inform policy and best practices. Limited research, knowledge and evidence is available about MNH outcomes in relation to the densities of the HWF delivering health services such as midwives, nurses and medical doctors. The aim of this study was to assess how HRH policies and strategies have influenced the trends of the maternal and newborn HWF densities, and the association between HWF densities, service coverage and health outcomes. We focused on stillbirths, LBW, preterm birth, neonatal and maternal mortality as outcomes as they contribute to the main burden of adverse outcomes. Moreover, the majority of these perinatal and maternal adversities are preventable if appropriate timely quality care is provided. [32] For health service coverage we applied two essential indicators – coverage of antenatal care of four or more visits (ANC4+) and SBA. The case study countries are Benin, Malawi, Tanzania and Uganda. These countries were selected to complement earlier research related to midwifery care providers’ competencies after pre-service training and intrapartum competencies conducted in the same countries. [10, 33] In addition, these countries are on the WHO’s “health workforce support and safeguards list, 2023. [34] Country inclusion in the list is based on a national density of doctors, nurses and midwives below the global median of 49 per 10 000 population and an universal health service coverage index below 55 (out of 100). [35] Table 1 provides density data of medical doctors nursing and midwifery personnel for each of the countries.

**Table 1.** Density of medical doctors, nursing and midwifery personnel

| Density by occupation<br>(per 10 000 population)<br>(latest available year) | Countries |  |  |  |
| --- | --- | --- | --- | --- |
|  | Benin | Malawi | Tanzania | Uganda |
| Medical doctors | 1.95<br>(2022) | 0.49<br>(2022) | 1.33<br>(2022) | 1.68<br>(2022) |
| Nursing personnel | 4.41<br>(2022) | 4.70<br>(2022) | 5.50<br>(2018) | 15.70<br>(2022) |
| Midwifery personnel | 1.54<br>(2022) | 0.30<br>(2022) | Not reported after 2006 | 6.98<br>(2022) |

Source: WHO National Health Workforce Account (NHHWA) Data Platform - Country profiles. December 2023 update. Accessed 03 January 2024.

This study aimed to answer the following research questions:

How have HRH policies and strategies influenced the changing trends of maternal and newborn HWF densities in Benin, Malawi, Tanzania and Uganda between 2010 to 2022?
What is the association between HWF densities, service coverage and perinatal and maternal health outcomes in Benin, Malawi, Tanzania and Uganda from 2010 to 2020?

## Methods

### Study design and setting

Various contextual factors, HRH policies and strategies and MNH health outcomes are present in Benin, Malawi, Tanzania and Uganda, and therefore an exploratory case-study policy process tracing approach was applied to each country’s health and policy data. Policy process tracing is a method that attempts to identify the underlying processes in order to explain outcomes “what mechanismic explanation accounts for the outcome?”. [36] The approach aims to determine whether an independent variable(s) really affects the dependent variable(s) and to what effect. The independent variables in the study (HWF densities by occupation) are regarded as causes of the change in the dependent variables (coverage of health services and MNH outcomes).

Two sets of data were collected and analysed to answer the research questions. In this case, data were required pertaining to policies and strategies relevant to HRH as well as data pertaining to HWF densities, service coverage and MNH outcomes.

We used the READ framework for our policy and strategy document analysis, a step-by-step guide for qualitative policy analysis. As a systematic approach, READ provides a framework for identifying appropriate documents and gleaning relevant information from them. The stages of READ are: 1) Ready your materials, 2) Extract data, 3) Analyse data and 4) Distil your findings. Each stage will be discussed as we outline data collection and data analysis methods. [37]

Lowess smoothing, which stands for locally weighted scatterplot smoothing, a variation of Lowess that uses a robust weighting scheme to reduce the impact of outliers was used for the comparative analysis. Lowess smooting is a visual method to fit smooth curves to the empirical data, without any specific assumptions, by using weighted linear regression. [38] This comparative analysis included five health outcome variables LBW, stillbirth (SBR), preterm birth (PTB), neonatal mortality (NMR) and maternal mortality (MMR), two service coverage variables (ANC4+ and SBA) and three maternal and newborn HWF densities (medical doctors, nursing, and midwifery personnel). All the empirical data used are quite variable in their structural patterns, data collection methods and availability of data points overtime. Hence, the key advantage in using the Lowess smoothing is that the fitted curve for any variable is obtained empirically rather than through stringent prior specifications about the nature of any structure that may exist within the data. The result is a smooth curve that follows the general trend of the data, but also adapts to local variations. [38]

### Data compilation and management

Under the READ framework we “Readied our materials”. This required us to set parameters in terms of the nature and volume of documents we wished to identify and analyse. This was achieved by defining the following criteria:

i. Topic: HRH policies and strategies related to maternal and newborn health.
ii. Dates of inclusion: We opted to include documents published from 2010 onwards to explore the policy environment. The year 2010 was also chosen as all our MNH health outcomes variables of interest have time trends data available from 2010 to 2020.
iii. Places to search for relevant documents available in the public domain: To obtain HRH policies and strategies we searched three different sources considered most pertinent: the Ministry of Health website for the four countries, WHO Global Sexual, Reproductive, Maternal, Newborn, Child and Adolescent Health (SRMNCAH) Policy Database (https://platform.who.int/data/maternal-newborn-child-adolescent-ageing/national-policies), and African Index Medicus. All sources were accessed between 01-06 January, 2024. The SRMNCAH Policy Database, a policy document repository, contains document shared by WHO Member States including national policies, strategies, laws, guidelines, and reports that are relevant to the areas of SRMNCAH obtained during rounds of the Global SRMNCAH Policy Survey. The African Index Medicus is an international database for African health literature implemented by WHO and African partners. The key words used were: human resources for health, health policies, health system, strategic plan and the country names.

Once relevant documents were identified using the above criteria, we moved onto the second stage of the READ framework to “Extract data”. This was achieved by using an Excel spreadsheet with each column representing a category of information we wished to extract and pertinent to the AAAQ framework. Documents were closely read by two reviewers (ABM and JW) with required information extracted and added to the spreadsheet (ABM and JW) (S1_Table).

Data relating to HWF density of nursing and midwifery personnel, and medical doctors were accessed and downloaded from the WHO National Health Workforce Account (NHWA) Data Platform (https://apps.who.int/nhwaportal/). The WHO NHHWA Data Platform hosts global HWF statistics including specific country profiles, key data on stock by occupational groups, age distribution and other key HWF indicators. Data were downloaded 03 January, 2024.

The country level data for PTB, LBW and MMR were retrieved from WHO’s Global Health Observatory (GHO) (https://www.who.int/data/gho) and the STB and NMR data from United Nations Children’s Fund (UNICEF) data website (http://data.unicef.org). Service coverage data, ANC4+ and SBA were also retrieved from the GHO. All data sets were downloaded 03 January, 2024 and are publicly available.

Maternal and perinatal definitions used in the study are described in Box 2.

S2_Text provides a brief summary of how the maternal and perinatal estimate data sets are generated.

#### Box 2.

**Maternal and perinatal definitions**

**Preterm birth:** less than 37 completed weeks (less than 259 days) of gestation.

**Low birthweight:** a birthweight less than 2500 g (up to and including 2499 g).

**Stillbirth:** the complete expulsion or extraction from a woman of a fetus, following its death prior to the complete expulsion or extraction, at 22 or more completed weeks of gestation. For international reporting it is recommended to report stillbirths of 28 or more completed weeks of gestation.

**Neonatal death:** a death during the first 28 completed days after live birth (days 0-27).

**Maternal mortality:** the death of a woman while pregnant or within 42 days of termination of pregnancy, irrespective of the duration and site of the pregnancy, from any cause related to or aggravated by the pregnancy or its management but not from unintentional or incidental causes.

Source: ICD-11 (https://icd.who.int/en).

## Data analysis

Once all relevant policy documents had been retrieved and pertinent information extracted and added to the Excel sheet, we followed step three of the READ framework, “Analyse data”, an iterative process of data synthesis by reviewing the content of the documents closely and considering their purpose. Following step four of the READ framework, we then “Distilled our findings”. This was achieved by mapping the relevant information from the policies and strategies to the AAAQ framework using the operational definitions to synthesise the data. Main categories were developed where possible for each aspect of the AAAQ framework. The documents were divided into two groups: i) specific HRH policy and strategic plans (overarching HRH policy/strategy) and ii) other policies and strategic plans considering HRH aligned to a broader health policy or health sector strategic plan. This was done as a very limited number of specific HRH policies and strategies are available for these countries and we wanted to explore how HRH was embedded in other policy and strategy documents which could provide critical information.

For the comparative analysis datasets (HWF densities, service coverage and outcomes) were merged (S3_Table) and imported into Stata 17.0 (StataCorp. 2023. Stata Statistical Software: Release 17. College Station, TX: StataCorp LLC.).

Instead of using a fixed neighbourhood size, Lowess smoothing adjusts the size of the neighbourhood based on the density of the data points in the scatterplot (xi, yi) where x are the years between 2010 to 2022 and y are the value of the variable of interest (five health outcome variables, two service coverage variables and the HWF densities by occupation). We generated individual country scatterplots with Lowess smoothing illustrating the relationship between i) HWF densities, LBW, PTB and ANC4+, ii) HWF densities, NMR, SRB, and SBA, and iii) HWF densities and MMR.

## Ethical consideration

All data used in the study are aggregated and publicly available therefore ethical approval was not required.

## Results

### Policy and strategy review

Twenty HRH policies and strategies are included in the analysis covering the period from 2010 to the most recent identified which was published in 2021. Three policies were identified from Benin, all of which fell in the category ii “other policies and strategic plans considering HRH” as no specific HRH policy and strategy was identified. From Malawi, five policies were included, one specific HRH strategy (draft) and four other policies and strategic plans. According to the literature it is documented that Malawi has an HRH Strategic Plan, 2018-2022 [39, 40] however, currently it is not available in the public domain. Two HRH strategies are included from Tanzania and three other policies and strategic plans. From Uganda one HRH strategy and six other policies and strategic plans were identified. S4_Fig shows the PRISMA flow diagram charting this process.

Benin is the only country which does not have an overarching HRH policy and strategy, whereas Malawi and Tanzania have developed two HRH policies/strategies. Uganda recently (2021) published a new strategy covering the period from 2020-2030 which is a follow on to the 15-year HRH Strategic Plan 2005-2020 which ended in June 2020. A detailed operational plan with targets and cost is only included in the Uganda HRH strategy 2020-2030 with 5-year rollout operational plan for 2020/21-2024/25. The HRH strategies from Malawi and Tanzania included strategic objectives and implementation framework but no concrete plan on how to achieve the objectives.

In addition to the overarching HRH policy and strategy each country has included aspects related to HRH in their health policy and/or health sector strategic plans.

All the documents have specific reference to maternal newborn and/or infant mortality and describe concerns about continuous high levels of maternal and newborn mortality that continue to fall behind national and global targets. Adverse outcomes such as preterm birth, LBW and stillbirth are not mentioned or considered in any of the documents.

In terms of the AAAQ framework, all HRH specific policies addressed availability of the HWF. All HRH policies (Malawi, Tanzania and Uganda) identified a need to focus on monitoring and planning for HRH, with all referring to retention of staff as a method to increase availability. Tanzania and Uganda also commented on the need to strengthen recruitment, with Uganda also noting the importance of reducing absenteeism as mechanisms for improving availability of staff. With regard to accessibility, all HRH specific policies commented on the need for an equitable distribution of HRH staff. Acceptability was poorly represented in policies for HRH with only the Ugandan policy mentioning the need to improve acceptability but providing no further detail in terms of what this means or how to achieve it. All HRH specific policies referred to quality of HRH in terms of improving pre-service training, as well as improving continuous professional development. To achieve the former, the Tanzanian policies discussed improving training curricula, strengthening the capabilities of teaching faculty and improving accreditation for training institutions. The Ugandan document also discussed improving training curricula as well as the need to improve clinical training, standardize exams, performance and manage staff throughout their working life (Table 2. AAAQ framework and human resources for health policies and strategic plans).

**Table 2.** Availability, Accessibility, Acceptability and Quality (AAAQ) framework and human resources for health policies and strategic plans

| Country | Title of policy/ strategic plan/year | Availability | Accessibility | Acceptability | Quality | Specific reference to MNH |
| --- | --- | --- | --- | --- | --- | --- |
| **Benin** | None identified |  |  |  |  |  |
| **Malawi** | Malawi Human Resources for Health Strategic Plan, 2018 – 2022. Not available in the public domain but have been referenced in the literature [39, 40] |  |  |  |  |  |
| **Malawi** | Human Resources for Health Strategic Plan 2012–2016. Draft [41] | Improve the capacity for HRH monitoring and planning.<br>Improve HRH retention. | Adequate and equitable distribution of HRH. |  | Strengthen HRH training and continued professional development.<br>Support capacity building agendas of health training institutions. | Reduction in maternal and newborn mortality. |
| **Tanzania** | Human Resource for Health and Social Welfare Strategic Plan 2014-2019 [42] | Improve the capacity for HRH monitoring and planning.<br>Improve HRH retention.<br>Strengthen recruitment. | Equitable distribution of HRH. |  | Strengthen HRH training and continued professional development (i.e. improve curricula for training programs and improve accreditation of training institutions). | Reduction in maternal and under five mortality. |
| **Tanzania** | National Human Resources for Health Strategy 2020-2025 [43] | Improve the capacity for HRH monitoring and planning.<br>Improve HRH retention.<br>Strengthen recruitment. | Equitable distribution of HRH.<br>HRH recruitment in low density areas. |  | Strengthen HRH training and continued professional development (i.e. improve curricula for training programmes, competence-based curriculum and faculty). | Reduction in maternal and newborn mortality. |
| **Uganda** | The Human Resources for Health Strategic Plan 2020-2030 [44] | Improve the capacity for HRH monitoring and planning.<br>Improve HRH retention.<br>Strengthen recruitment.<br>Reduce absenteeism. | Adequate and equitable distribution of HRH. | Improving acceptability of workforce is mentioned, but no plans or examples of how to do this are given. | Strengthen HRH training and continued professional development (i.e. improve curricula for training programmes, improved clinical training, standardized examination).<br>Maintain regular performance reviews and appraisals based on performance plans and contracts.<br>Strengthen professional associations as a mechanism<br>for HWF empowerment. | Reduction in maternal and newborn mortality. |

For policies and strategies not specific to HRH, all documents from Benin, Malawi and Tanzania, and four out of the six documents from Uganda addressed availability. To improve availability, a variety of strategies were suggested including strengthening HRH planning (Benin, Malawi, Tanzania and Uganda), ensuring an appropriate supply of staff (Benin, Malawi, Tanzania and Uganda), focusing on recruitment (Benin, Malawi and Uganda) improving retention (Benin, Malawi, Tanzania and Uganda) and reducing absenteeism (Uganda). With regard to accessibility, all documents from Benin, Malawi, Tanzania and one of the six documents from Uganda mentioned improving HRH accessibility. This was discussed in terms of equitable distribution of HRH (Benin, Malawi, Tanzania and Uganda), provide health facilities with appropriate equipment (Benin), appropriate referral systems (Malawi, Tanzania and Uganda), improved access to emergency transport (Malawi), and addressing financial barriers in accessing care (Malawi and Tanzania).

As with the HRH specific policies, the concept of HRH acceptability was poorly represented in other policies and strategies with no mention of the concept in the Benin policies and strategies, and only one of the four documents from Malawi addressing it. Tanzania and Uganda mentioned acceptability more consistently with all Tanzanian documents and four of the six Ugandan documents identifying the need to improve HRH acceptability. Strategies suggested for achieving this included client-centredness and being considerate of people’s personal circumstances, meeting the expectations of the population, addressing community needs, people-centred respectful care with a human rights-based approach, improving professional standards and accountability towards clients, and client charters. For each country, all documents made recommendations for improving HRH quality. Strategies included improving training (Benin, Malawi and Tanzania) and continuous professional development (Benin, Malawi and Tanzania), improving staff motivation (Benin and Malawi), accreditation of training institutions (Malawi and Uganda), performance management of staff (Tanzania and Uganda), strengthening regulatory bodies (Uganda), and ensuring staff have access to and follow guidelines and protocols (Uganda). Details provided in S5_Table.

From the review it is also observed that none of the policies and strategies mentioned the SDG target 3.c and indicator 3.c.1. related to the HWF. [2]

### Comparative analysis – health workforce densities, maternal and newborn services and health outcomes

The patterns of change in the perinatal health outcomes – PTB, LBW, NMR and SBR – appear to be insensitive to the fluctuations in HWF densities and HRH policies and strategies implemented since 2010 indicated by the stagnation or minimal reduction observed across all the perinatal outcomes.

A reduction in MMR is observed in all the countries, even with no change in the midwifery density as observed in Benin and Malawi, whereas in Uganda the midwifery density has steadily increased since 2013. In Tanzania nursing and midwifery personnel is most often dual trained combining nursing and midwifery [10] and reported here under nursing personnel.

The nursing density increased from 2014 to 2017 with slight decrease in 2018 and after no data have been reported.

Limited data are available for coverage of ANC4+ but from the reported data it becomes apparent that PTB and LBW rates are not reactive to levels and fluctuations in ANC4+ coverage. A similar pattern is seen for NMR and SBR as no changes in these outcomes are observed even though all countries have reported increase in SBA.

(Fig 1a, 1b, 1c, 2a, 2b, 2c, 3a, 3b, 3c, 4a, 4b and 4c)

**Fig 1a.**
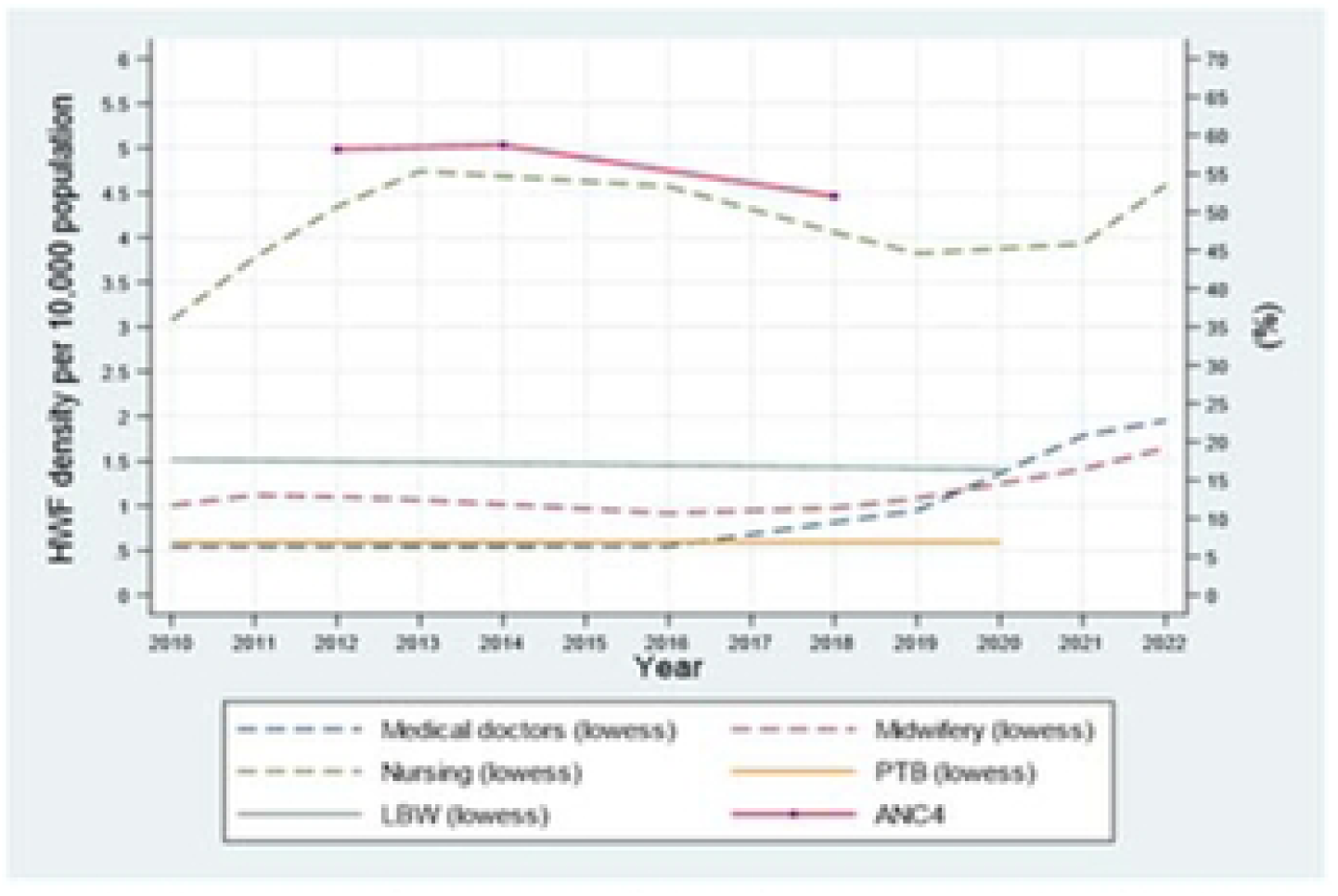
Benin HWF densities in relationship to LWB, PTB and ANC4.

**Fig lb.**
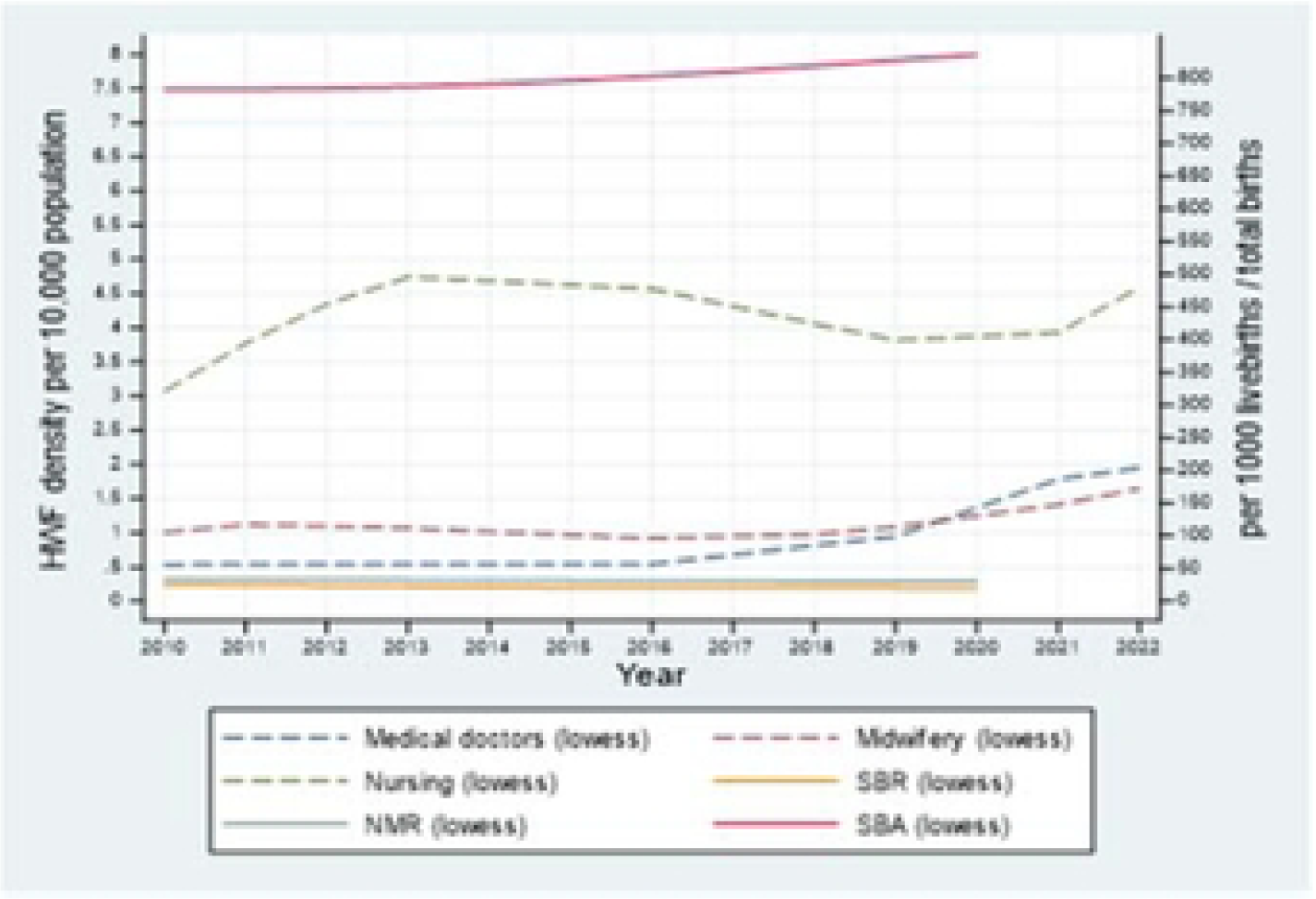
Benin HWF densities in relationship to NMR, SBR and SBA.

**Fig lc.**
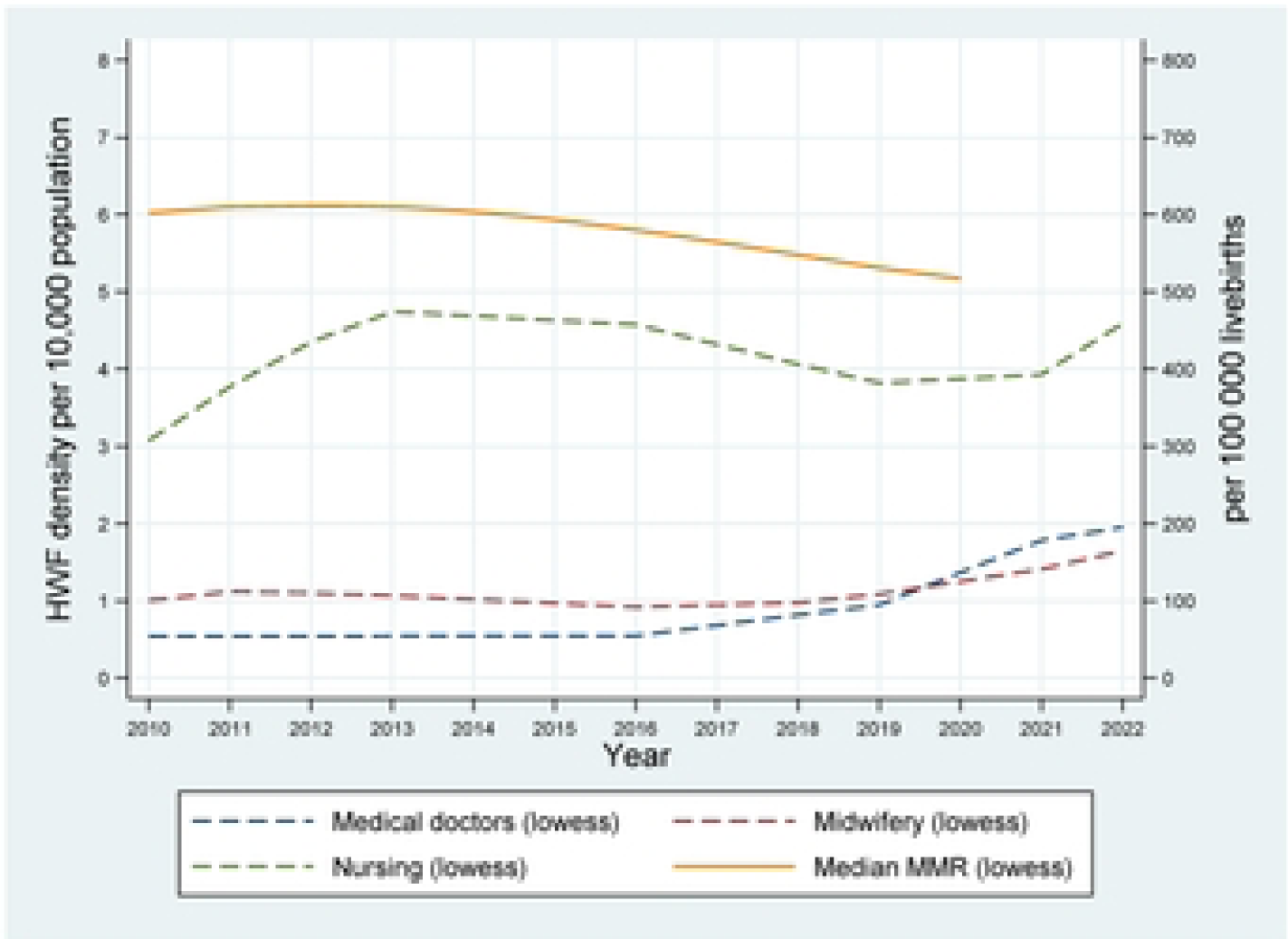
Benin HWF densities in relationship to MMR.

**Fig 2a.**
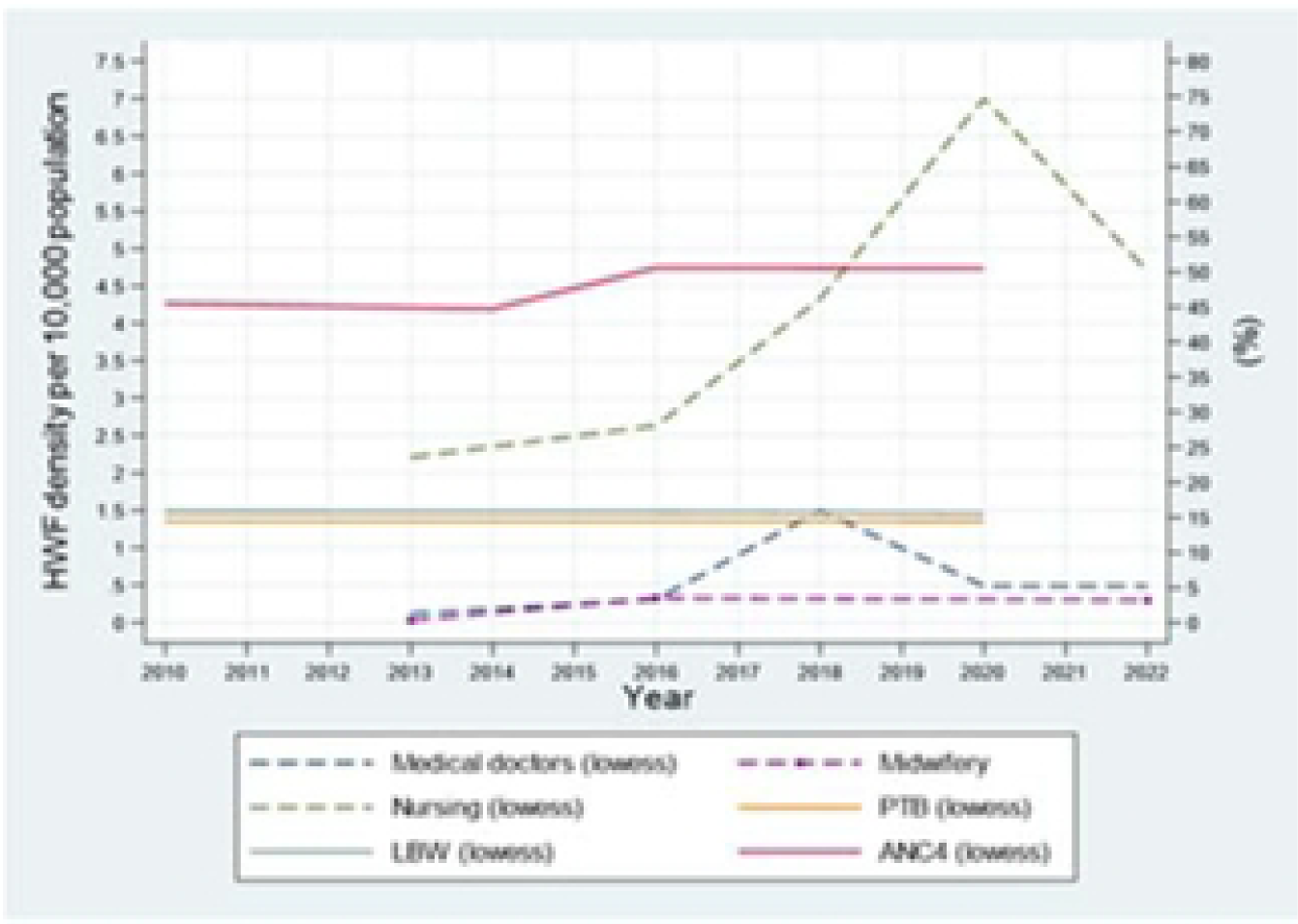
Malawi HWF densities in relationship to LWB, PTB and ANC4.

**Fig 2b.**
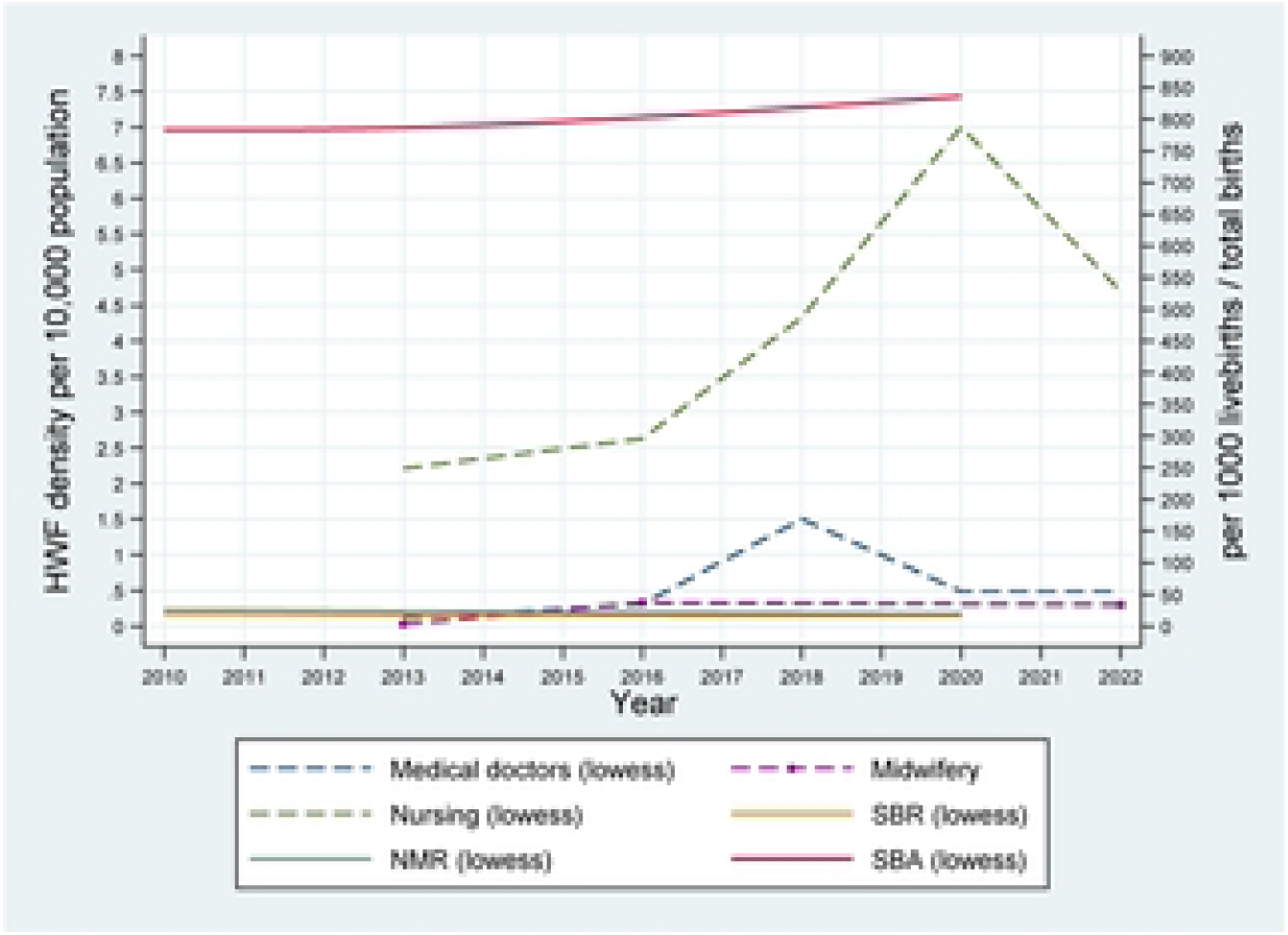
Malawi HWF densities in relationship to NMR, SBR and SBA.

**Fig 2c.**
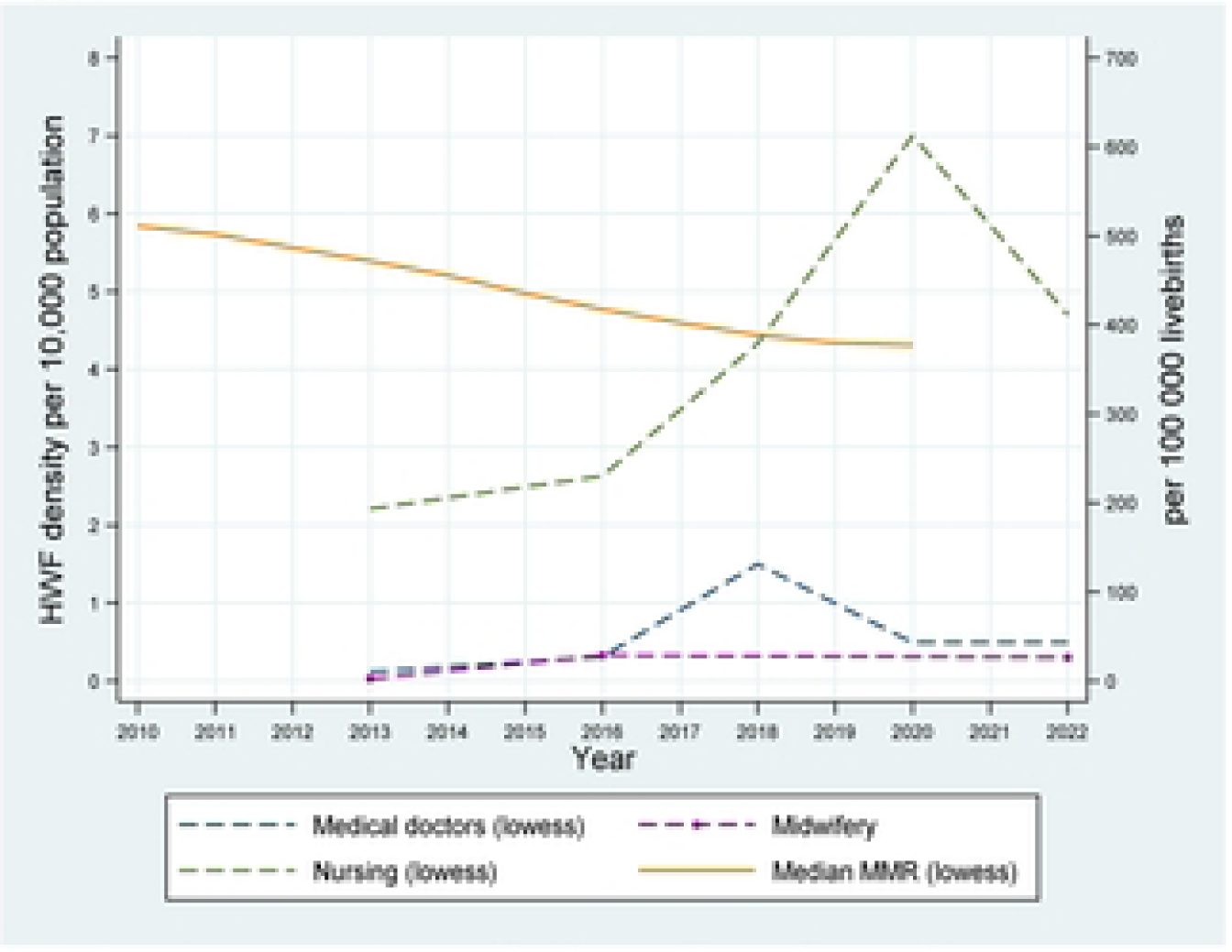
Malawi HWF densities in relationship to MMR.

**Fig 3a.**
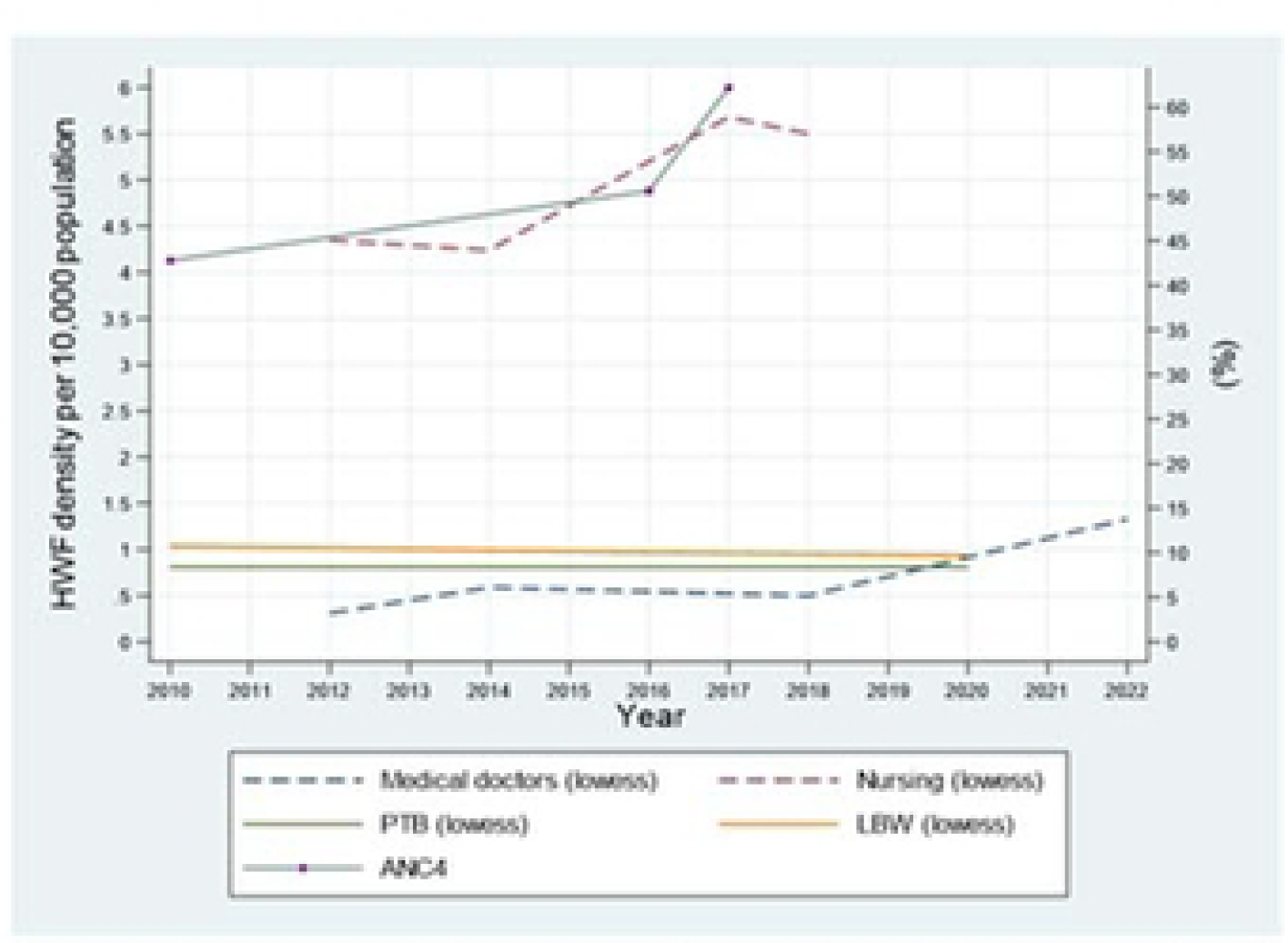
Tanzania HWF densities in relationship to LWB, PYB and ANC4.

**Fig 3b.**
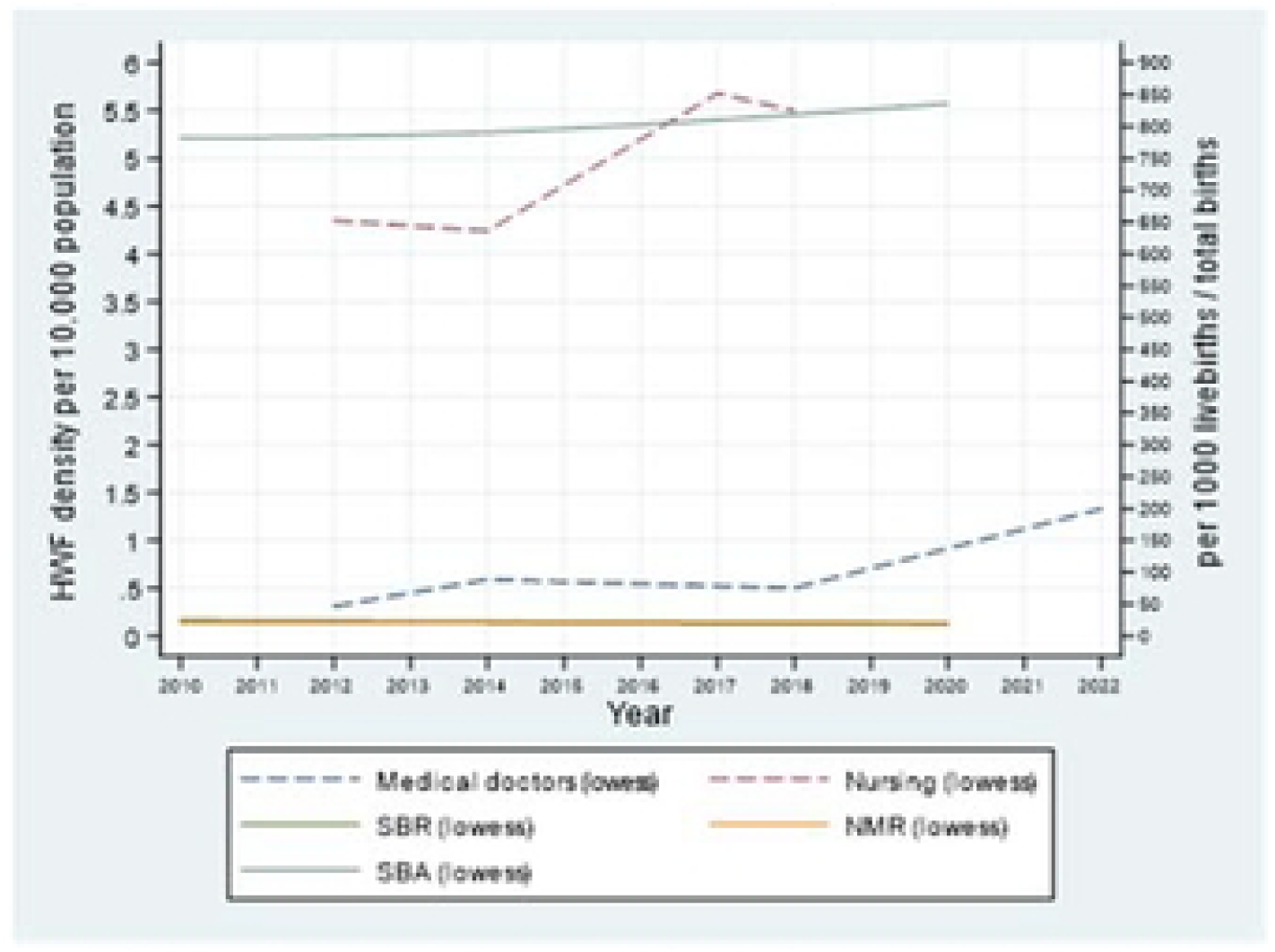
Tanzania HWF densities in relationship to NMR, SBR and SBA.

**Fig 3c.**
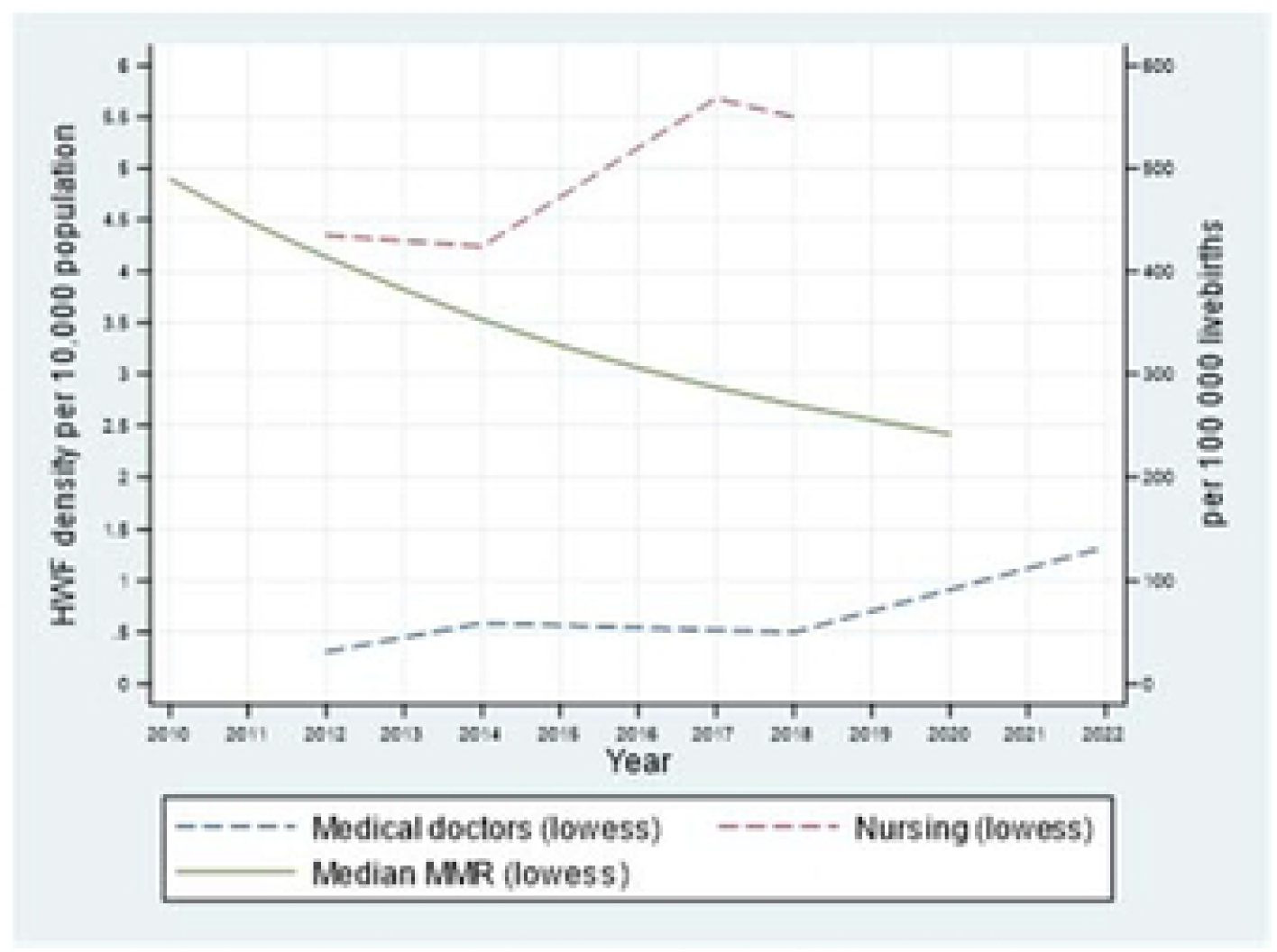
Tanzania HWF densities in relationship to MMR.

**Fig 4a.**
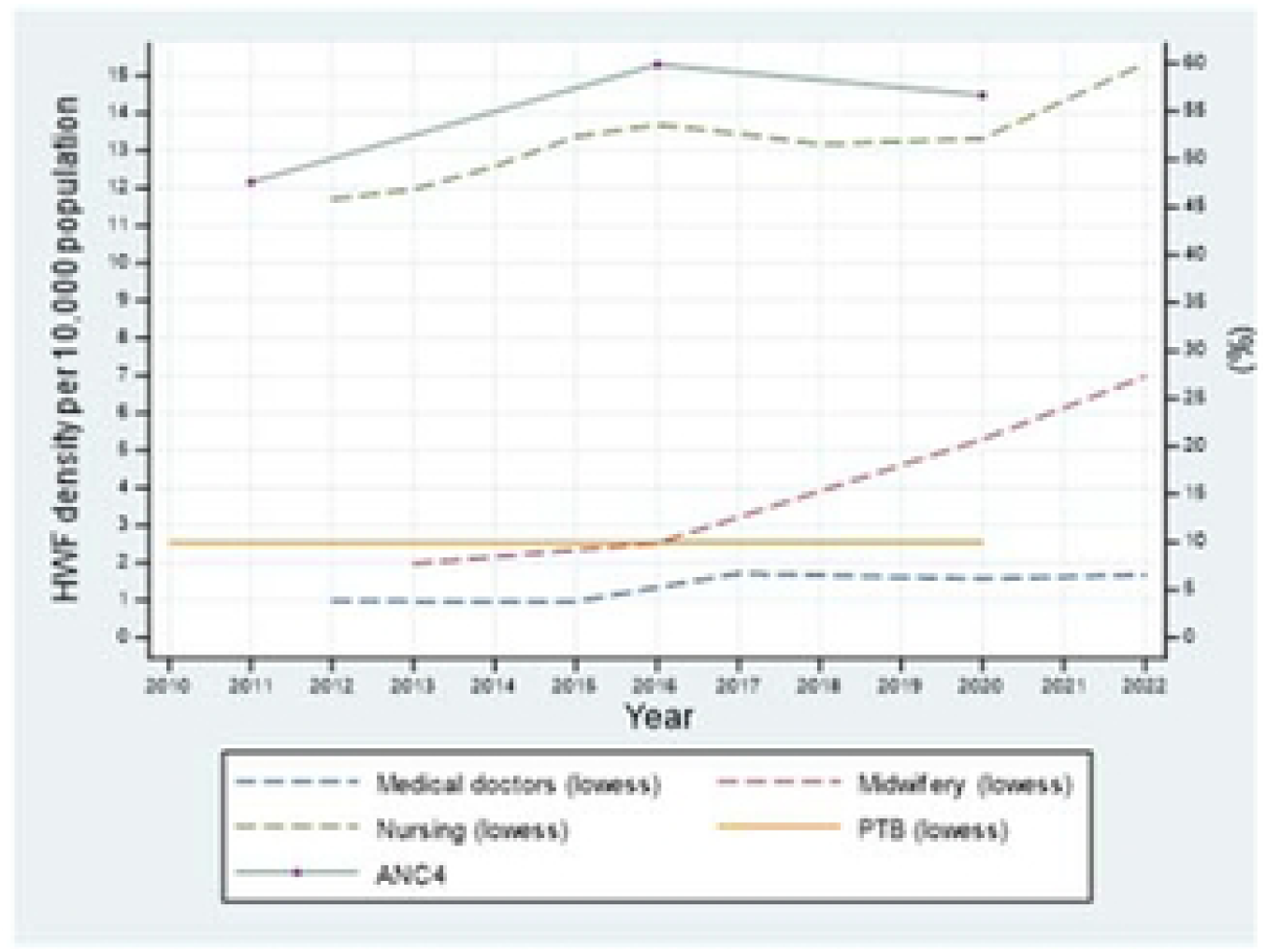
Uganda HWF densities in relationship to PTB and ANC4.

**Fig 4b.**
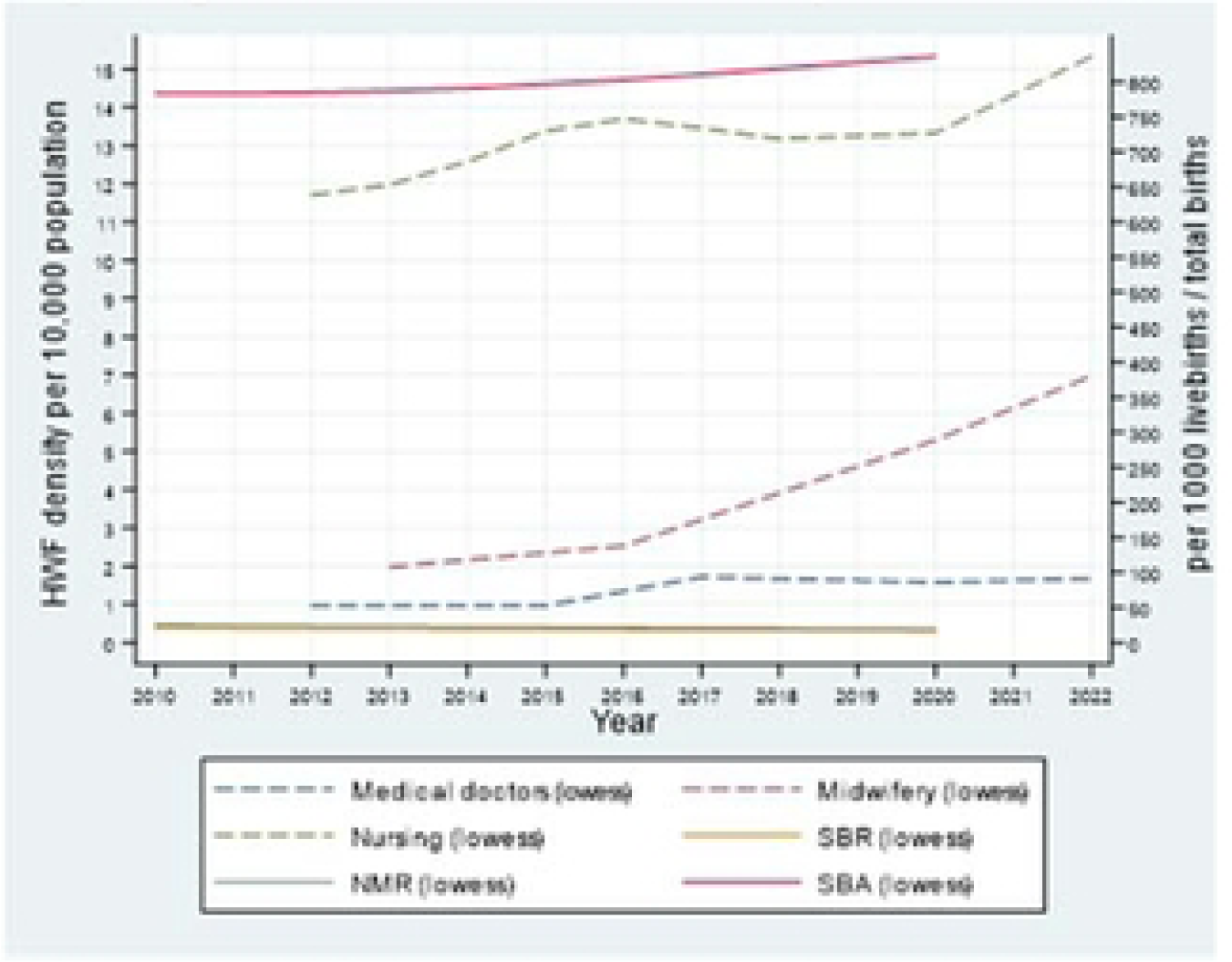
Uganda HWF densities in relationship to NMR. SBR and SBA.

**Fig 4c.**
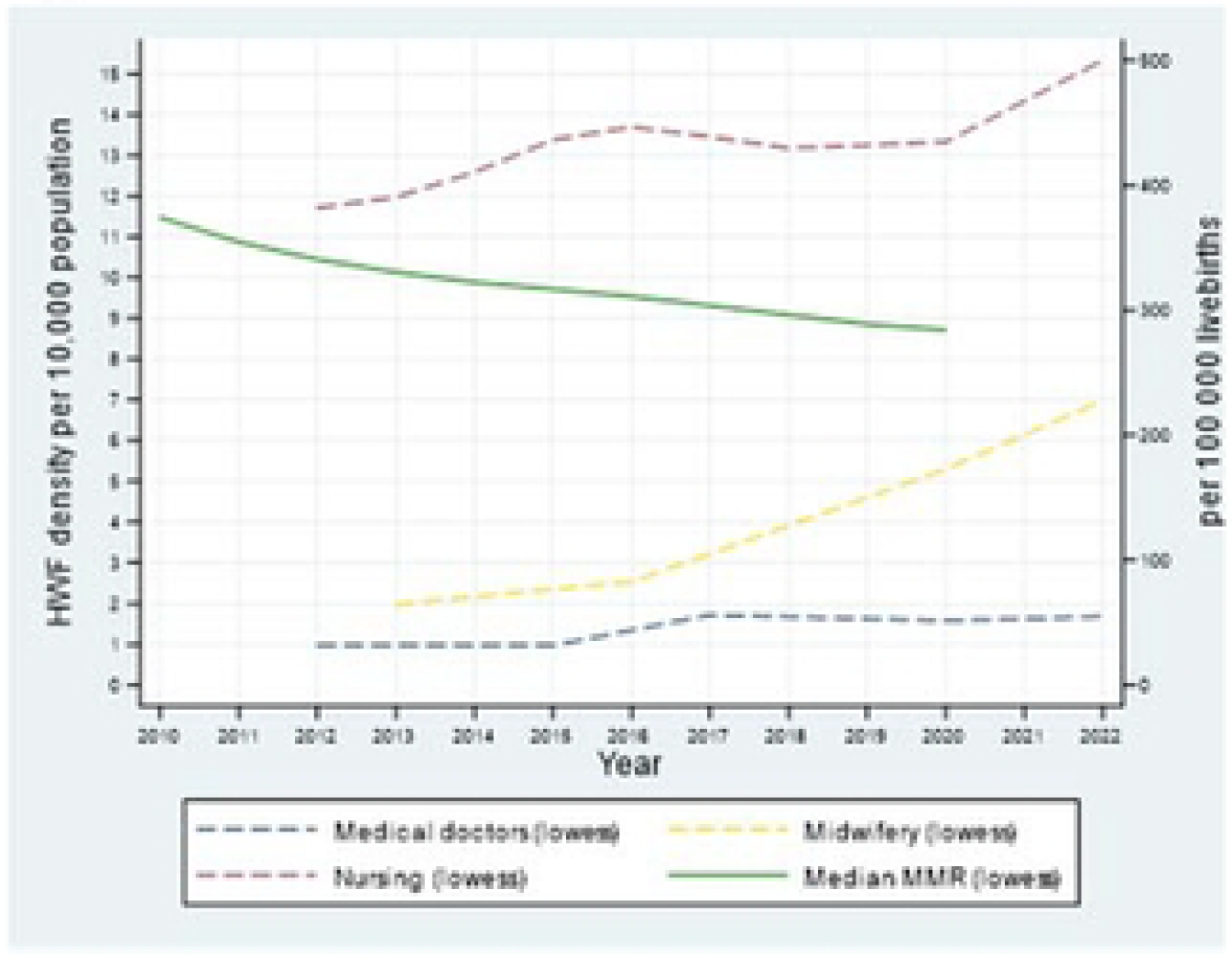
Uganda HWF densities in relationship to MMR.

### Benin

In Benin the “National Health Development Plan 2009-2018” [45] preceded an increase in the nursing density (from 3.34 in 2010 to 6.42 in 2016 per 10 000 population) with no change in medical doctors and midwifery densities. More recent policies, the “Operational plan to reduce maternal and neonatal mortality in Benin 2018-2022” [46] and the “National Health Policy 2018-2030” [47] emphasized the need to increase the number of health care professionals, especially obstetricians/gynaecologists, paediatricians and midwives. Moreover, the report stressed the need for more competent midwives noting that HRH were lacking in relation to MNH care and services. It appears these policies have been reflected as both medical doctors and midwifery densities increased from 2018. At the same time the nursing density decreased but increased again in 2021 (Fig 1a, 1b and 1c).

### Malawi

The “Human Resources for Health Strategic Plan 2012-2016” in Malawi [41] generated an increase in the densities of medical doctors and nursing with no change in midwifery density. The strategic plan emphasises the need for improved HRH capacity, better monitoring and planning as well as the need to strengthen pre-and in-service training including enhancement of training institutions capacity. The “Human Resources for Health Strategic Plan 2012-2016” was supported by the “Health Sector Strategic Plan 2011-2016. Moving Towards Equity and Quality” [48] which stressed similar aspects where enhancements were required but also highlighted the need for pre-service training curricula revisions to reflect new evidence and national standards. A peak in medical doctor density occurred between 2016 to 2018 (from 0.31 to 1.5 per 10 000 population) which then decreased in 2020 (0.49 per 10 000 population) to almost the same level before the observed increase in 2018. A similar peak, just few years later in 2020 occurred for nursing density with an increase from 4.33 in 2018 to 7.0 in 2020 per 10 000 population. After the 2020 peak the 2022 density was back at the 2018 level (4.33 per 10 000 population). During this period two strategies were in place the “Health Sector Strategic Plan II 2017-2022. Towards Universal Health Coverage” [49] and the “National Health Policy - Towards Universal Health Coverage 2018-2030” [50] both of which stressed the need for improved HRH capacity, retention, a more equitable distribution of HRH and better coordination of pre-and in-service training to enhance staff competence. Changes in trends and these peaks of HWF densities may reflect changes in data reporting mechanisms. A true understanding of the fluctuations in the HWF densities therefore requires thorough investigation into reporting mechanisms and recruitment patterns (Fig 2a, 2b and 2c).

### Tanzania

No data has been reported on density of nursing in Tanzania since 2018. However, a slight increase from 2012 to 2018 (4.35 to 5.5 per 10 000 population) occurred. Density data are available for medical doctors with only a 23% increase per 10 000 population observed from 2012 to 2022 (0.31 in 2012 and 1.33 in 2022 per 10 000 population). Two HRH strategies, the “Human Resources for Health and Social Welfare Strategic Plan 2014-2019” [42] and the “National Human Resources for Health Strategy 2020-2025” [43] are the basis for the HRH strengthening and programme development. Emphasis in both strategies is to improve HRH capacity, planning, monitoring and enhanced training however, from the current available data it is not possible to distil how these strategies have affected the nursing personnel density (Fig 3a, 3b and 3c).

### Uganda

Both nursing and midwifery densities have continued to increase since 2012 reaching a level of 15.7 nursing per 10 000 and 7.0 midwifery per 10 000 population in 2022. The density of medical doctors has only slightly increased in the same period with a density of 1.68 per 10 000 population in 2022. The “Second National Health Policy (NHP II) 2010” [51] may have been the impetus for strengthening the nursing and midwifery with several other strategies supporting this [44, 52–56], each giving prominence to the need for HRH with the appropriate capacity, skill mix, training, accreditation and a focus on reducing absenteeism (Fig 4a, 4b and 4c).

Placeholder: Please insert Fig 1a, 1b, 1c, 2a, 2b, 2c and Fig 3a, 3b, 3c, 4a, 4b, 4c

## Discussion

Findings from the review of strategies and policies revealed a number of suggested approaches to improve the availability of the HWF including recruitment, retention and tackling absenteeism. Recruitment of health care staff would appear to be an obvious solution to HWF shortages however, reasons exist as to why this may not be possible. National government budgets for healthcare may not align with health care service demands, leading to an inability to recruit the required HWF. Commentary from Malawi suggests this is a real issue. Muula (2023) posits that Malawi produces hundreds of health care professional graduates annually, who if employed would resolve HRH shortages. [57] Evidence suggests that improving retention of the current HWF is complex. Retention is associated with working conditions, availability of equipment and resources, ability to provide quality care, salary, opportunities for career progression, housing availability and quality (where housing is provided) as well as the feeling of being valued by colleagues. [58–60] Addressing these issues is multifaceted and requires significant investment from national government.

Reducing absenteeism as a means to improve HRH availability was proposed in four of Uganda’s policies and strategies. However, absenteeism was not mentioned in any policy documents from Benin, Malawi and Tanzania. Whilst acknowledging the complexity of HRH absenteeism, Ackers et al. [59, 61], go as far as saying that failure of doctors to present for work is one of the causes of maternal mortality in Uganda. Research by Zhang et al. (2021) in Uganda found a 15% absenteeism prevalence on monitored days, but this rate was higher, 42% in lower-level health clinics. [62] Such absenteeism not only affects the availability of the HWF, but also the accessibility and acceptability of HRH, with absenteeism associated with lower rates of health care utilization at lower-level facilities. Whilst studies call for a crackdown on absenteeism [59, 62], Di Giorgio et al. (2020) [63] highlight that reducing absenteeism without addressing gaps in health care worker competence would only modestly increase the average care readiness that meets minimum quality standards. Tackling HWF absenteeism requires a greater understanding of its causes, which may include working conditions and remuneration. Addressing these may improve availability of the HRH workforce in SSA.

Our review of the strategies and policies highlighted recommendations for an equitable distribution of the health care workers between rural and urban areas, as well as reductions in out-of-pocket expenditure for patients as a means of improving patient accessibility to the HWF. The unequal distribution of health care facilities and resources, including the HWF, is not a new phenomenon with these disparities widely reported in the literature. [1, 64] Similarly, suggestions for tackling these issues are not new, and have been consistently recommended in various reports and strategies for several years. [65–68] Realizing the ambition of an accessible HWF will require significant financial commitment from government to ensure health clinics and hospitals that are equipped with adequate health workers and resources are made readily available in the most needed areas.

Notably, few strategies referred to improving the acceptability of HRH. This is somewhat surprising, as the literature pertaining to the importance of respectful maternity care is abundant. [69–71] Research from all four study countries indicates respectful care in the maternity setting is lacking. [72–76] Tackling disrespect and abuse in the health care setting, particularly the maternity setting, is an essential component of improving acceptability of HRH.

Improving the quality of the HWF competence is imperative for better and desirable health outcomes and if quality of care is to be enhanced. The documents reviewed in this study heavily focused on the need to improve the quality of pre-service training for health care professionals as well as continuous education as a means of improving the quality of the HWF. These recommendations are valid given current evidence which suggests deficiencies in pre-service and in-service training of health care professionals contributes to a lack of a skilled HWF. [10, 77–79]

Many of the policies repeat themselves in terms of issues and solutions to HRH issues, with no clear evidence of change between strategies. Moreover, the strategies do not provide clear guidance on how to achieve change.

The HWF densities and perinatal outcomes comparative analysis clearly shows that the changing trends in HWF densities have not ensured any clear changes on the trends of PTB, LWB, NMR and SBR rates as no improvement in these outcomes have been achieved between 2010 to 2020. It was also noted in the policy review that PTB, LBW and SBR outcomes were not stressed or mentioned in any of the policies and strategies as a matter of priority and/or concern.

Current evidence indicates that increased rates of SBA have not been met with proportional decreases in maternal and neonatal mortality. [32] This demonstrates that access to care alone is not enough to improve health outcomes. Evidence-based quality care is essential. Poor quality care has been identified as a driver for excess mortality across a range of health care disciplines. [80]

## Limitations

The countries in this study may face challenges in reporting their HWF density data due to lack of comprehensive reporting systems and the ability to correctly apply the International Standard Classification of Occupations – ISCO-08 codes [81], which may be reflected in current data availability and accuracy. We acknowledge that the patterns of change in HWF densities are not solely influenced by national strategic plans and policies as many other factors will influence how these policies and strategies are implemented. Such factors include the economic climate, political will, pandemics, HRH in and-out migration and HRH management, all of which vary pending on the local context. Furthermore, a number of the strategies and policies reviewed in this article are recently published, thus may not yet have been fully implemented.

The comparative analysis only covers a relative short time period where a longer time period may have elucidated more information. However, the period was purposely selected as data for preterm birth were only available from 2010. The comparative analysis used aggregated data which may not represent the potential differences within the countries.

With regard to the visual comparisons conducted using Lowess smoothing, there was a certain degree of sensitivity observed given the scarcity of empirical data (for example for ANC4+) and here the choice was made to display the original data values as a simple scatter over the time period of comparison.

## Conclusion

Applying a policy tracing approach against a backdrop of changes in the trends of HWF and MNH services and outcomes over time is an informative method of improving the understanding of data for policy and practice. The Lowess visual method has shown the nature of the progress, if any, akin to the HRH influential policies. However, there are no observable improvements in perinatal outcomes relative to improvement in HWF availability measured by the densities, which would imply that other deterministic inputs govern perinatal outcomes at the service delivery level. The comparative analysis presented in this paper demonstrate the advantage of using Lowess-enhanced scatterplots that can tap on relatively complex relationships that could easily be overlooked with traditional statistical modelling procedures.

Tackling the issues that influence the availability, accessibility, acceptability, and quality of the HWF in Benin, Malawi, Tanzania and Uganda is no easy task. Powerful commitment at the highest-level, sustainable investment and agility in policy-decisions from national governments must be regularly monitored and reassessed to influence those four critical dimensions. Without these efforts, progress in these four countries towards women and newborn human rights to health will be inadequate and less likely the buildup of success towards UHC and the other health-related SDGs.

## List of abbreviations

AAAQ: Availability, Accessibility, Acceptability and Quality
ANC4+: antenatal care of four or more visits
CrI: credible interval
LBW: low birthweight
GHO: Global Health Observatory
MMR: maternal mortality ratio
MNH: maternal and newborn health
HRH: human resources for health
HWF: health workforce
NHWA: National Health Workforce Account
NMR: neonatal mortality rate
PTB: preterm birth
READ: Ready your materials, Extract data, Analyse data and Distil your findings
SBR: stillbirth
SDG: Sustainable Development Goals
SRMNCAH: sexual, reproductive, maternal, newborn, child and adolescent health
SSA: Sub-Saharan Africa
UHC: universal health coverage
UNICEF: United Nations Children’s Fund
WHO: World Health Organization

## Acknowledgments

None

## Authors’ contributions

Conceptualization: Ann-Beth Moller.

Data curation: Ann-Beth Moller and Joanne Welsh.

Formal analysis: Ann-Beth Moller, Joanne Welsh and Amani Siyam.

Funding acquisition: None.

Methodology: Ann-Beth Moller, Joanne Welsh and Amani Siyam.

Supervision: Max Petzold.

Writing original draft: Ann-Beth Moller.

Writing – review and editing: Ann-Beth Moller, Joanne Welsh, Amani Siyam and Max Petzold.

## Funding

The authors received no specific funding for this work.

## Competing interests

The authors have declared that no competing interests exist.

## Data availability statement

All original data are publicly available.

## Notes

### Competing Interest Statement

The authors have declared no competing interest.

